# Reusing Blood Samples from a Hospital-based Cohort to Measure Apixaban Plasma Concentrations

**DOI:** 10.64898/2026.04.07.26350322

**Authors:** Katherine T. Murray, Daniel V. Fabbri, Jeffrey S. Annis, Cynthia R. Clark, Jill M. Pulley, Evan L. Brittain, David Gailani

## Abstract

In the management of atrial fibrillation, the most frequently prescribed oral anticoagulant is apixaban, given at a fixed dose of 5mg BID. Apixaban is predominantly metabolized by cytochrome P4503A4 (CYP3A4) and is also a substrate for the drug efflux transporter P-glycoprotein (P-gp). In nearly 300,000 Medicare patients with AF receiving apixaban, we previously showed that concomitant therapy with drugs that inhibit both CYP3A4 and P-gp, specifically amiodarone or diltiazem, significantly increased serious bleeding that caused hospitalization and/or death. We hypothesized that this adverse effect was mediated by an increase in apixaban plasma concentrations caused by concomitant therapy that reduced drug elimination. Utilizing left-over samples obtained from clinically indicated blood draws that would typically be discarded, the Vanderbilt University Medical Center biobank BioVU contains >353,000 samples linked to de-identified electronic medical records (EMRs), with both DNA and plasma harvested. Of 35 samples drawn from patients taking apixaban 5mg BID, 5 were identified to be drawn from patients concomitantly taking drugs inhibiting both CYP3A4 and P-gp. Using a chromogenic anti-Xa assay, we found that plasma concentrations of apixaban were significantly higher (347±64 ng/mL; mean±SEM) for patients receiving concomitant CYP3A4/P-gp-inhibiting drugs compared to those not treated with these drugs (166±67 ng/mL; P=0.025, Mann Whitney). There were no differences between the 2 patient groups with respect to age, weight, or serum creatinine. The results of this pilot study provide preliminary data to support our hypothesis, and they demonstrate the practicality of obtaining pharmacokinetic data from a large cohort of plasma samples linked to deidentified EMRs. This approach could be used to define the role of apixaban levels in high-risk clinical scenarios and to better understand the relationship between drug levels and bleeding risk.

## Introduction

Atrial fibrillation (AF) is the most common sustained cardiac arrhythmia, and it increases the risk of stroke and thromboembolic events five-fold. In the management of AF, direct oral anticoagulants (DOACs) are first-line therapy for most patients, and the most frequently prescribed DOAC is apixaban.^1^ A therapeutic range for apixaban, or the plasma concentrations associated with bleeding events, have not been previously established. Thus, the drug is prescribed at a fixed dose of 5mg BID for most patients, and measurement of plasma concentrations to guide therapy is not recommended.^2^ However, there is increasing evidence that apixaban plasma concentrations in AF patients in clinical practice are higher than those documented in clinical trials,^3^ and that they can correlate with clinical outcomes.^4^ Moreover, for a given dose, there can be marked interpatient variability in plasma concentrations. Apixaban is predominantly metabolized by cytochrome P4503A4 (CYP3A4), and it is also a substrate for the drug efflux transporter P-glycoprotein (P-gp). Co-administration of drugs that inhibit the function of CYP3A4 or P-gp can increase plasma concentrations that could potentially lead to bleeding events.^2^ In nearly 300,000 Medicare patients with AF receiving apixaban, we have previously shown that concomitant therapy with drugs that inhibit both CYP3A4 and P-gp, specifically amiodarone or diltiazem, significantly increased serious bleeding that caused hospitalization and/or death.^5, 6^ We hypothesized that this adverse effect was mediated by an increase in apixaban plasma concentrations caused by concomitant therapy that reduced drug elimination.

## Methods

The DNA biobank at Vanderbilt University Medical Center, known as BioVU, is one of the world’s largest genomic repositories with >353,000 samples linked to de-identified electronic medical records (EMRs), with patient data spanning more than 15 years. Samples obtained from clinically indicated blood draws that would typically be discarded after several days are harvested and currently processed to bank both DNA and plasma.^7^ Of approximately 1000 recently-harvested plasma samples, 35 were drawn from patients taking apixaban 5mg BID. Blood samples were drawn regardless of timing relative to the previous apixaban dose. Using AI-based chart extraction (BRIM Analytics),^8^ and manual review of de-identified EMRs, we identified 5 of 35 samples from patients concomitantly taking drugs that are strong or moderate inhibitors of CYP3A4 that also inhibit P-gp (www.fda.gov; amiodarone in 4 and verapamil in 1). Plasma concentrations of apixaban were measured using a chromogenic anti-Xa assay (Stago). This report adheres to the SQuIRE 2.0 checklist (https://www.equator-network.org/reporting-guidelines/squire/).

## Results

The **Figure** illustrates the results for all patients taking apixaban 5mg BID at the time of blood draw. For those patients who were also receiving concomitant CYP3A4/P-gp-inhibiting drugs (Apix 5mg [+]), plasma concentrations of apixaban were significantly higher (347±64 ng/mL; mean±SEM) compared to those who were not being treated with these drugs (Apix 5mg [-]:166±67 ng/mL; P=0.025, Mann Whitney). It is well recognized that other demographic factors can influence apixaban plasma concentrations. However, there were no significant differences between patients taking CYP3A4/P-gp-inhibiting drugs and those not taking such drugs with respect to age (59.8±3 vs 62.8±2 years, respectively), weight (89.5±7 vs 97.7±4 years), or serum creatinine (1.2±0.1 vs 1.3±0.1 mg/dL).

**Figure.**
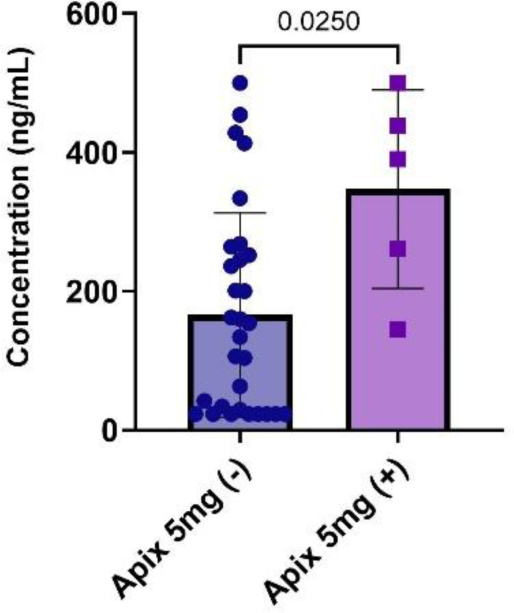

## Discussion

Previous studies to investigate the effect of drugs such as amiodarone on apixaban plasma concentrations have provided conflicting results.^9, 10^ The results of this pilot study provide preliminary data to support our hypothesis, and they demonstrate the practicality and potential value of a pragmatic approach to obtain real-world pharmacokinetic data from a large cohort of plasma samples linked to deidentified EMRs. This approach could be used to define the role of apixaban plasma concentration measurements in high-risk clinical scenarios to improve patient safety and to better understand the relationship between drug levels and bleeding risk.

## Data Availability

All data produced in the present study are available upon reasonable request to the authors

